# The epidemiology of chikungunya virus in Brazil and potential impact of vaccines

**DOI:** 10.1101/2025.07.08.25331112

**Authors:** Oscar Cortes-Azuero, Megan O’Driscoll, Gabriel Ribeiro dos Santos, Ronaldo de Jesus, Shirlene T. S. de Lima, Danny Scarponi, Christinah Mukandavire, Arminder Deol, Moritz U G Kraemer, William M. de Souza, Henrik Salje

## Abstract

The first chikungunya virus (CHIKV) vaccine is now licensed in Brazil, the country that reports most cases globally, however, the optimal use of the vaccine remains unclear due to a poor understanding of CHIKV epidemiology and population immunity. We quantified the annual CHIKV infection and disease burden since 2014 in each of the 27 Federal Units of Brazil using a mathematical model that integrated serological surveys (N=12), confirmed chikungunya cases (N=488,234) and chikungunya deaths (N=1,719) reported between January 2014 and September 2024. Using this base, we estimated the potential impact of a vaccine over the period 2025-2029 had the population been vaccinated before the 2025 season, evaluating different rollout strategies. We found that 18.3% (95%CrI: 16.5-20.3) of the population has been infected since 2014, with the highest risk concentrated in the Northeast and Southeast. Overall, 1.13% (95%CrI: 1.07-1.19) of infections were detected by surveillance systems, with an increasing probability of symptoms with age and greater risk of symptoms in females. Vaccinating 40% of the population aged >12y (73 million doses), and assuming a vaccine efficacy of 70% against infection and 95% against disease, would avert up to 1.6 million (95%CrI: 0.5-3) cases and 198 (95%CrI: 61-359) deaths over the next five years. Despite widespread circulation, most of the country remains vulnerable to infection. CHIKV vaccination has the potential to substantially reduce disease burden.

## Introduction

Chikungunya virus (CHIKV) is an arbovirus primarily transmitted by *Aedes aegypti* and *Aedes albopictus* mosquitoes. It is estimated to cause 35 million infections annually and over 3,000 deaths ^1,2^. The first CHIKV vaccines, IXCHIQ (formerly VLA1553) and VIMKUNYA (formerly PXVX0317), were licensed in the US and the European Union between 2023 and 2025 through accelerated pathways, however, their potential impact remains unclear ^3,4^.

A key barrier to vaccine implementation has been a poor understanding of the underlying epidemiology, and age- and sex-specific patterns of disease and death. A more detailed understanding of how the burden of disease is distributed within a country and across age groups is necessary to optimise vaccine policy. Characterizing levels of previous exposure from historic circulation is also an important consideration as they may limit the impact of the vaccine.

In most affected countries, CHIKV circulates epidemically, with infrequent large outbreaks followed by several years of few or no cases^1^. However, a few countries across three continents, including India, Kenya and Brazil, experience endemic transmission with cases reported annually^1^. Brazil reports the most chikungunya cases globally, with over 1.4 million suspected cases since the first autochthonous cases in 2014^2^. Nonetheless, it remains unclear how many undetected infections have occurred since its introduction. Brazil is geographically diverse with substantial heterogeneity in infection risk across the country^5^ and has recently licensed IXCHIQ vaccine. Therefore, understanding which regions to prioritise and the likely impact of vaccines will be an important step to data-driven vaccine strategy.

To understand the underlying burden of chikungunya, we can use the wealth of information collected through Brazil’s surveillance systems to reconstruct the history of CHIKV circulation. In particular, we combined the distribution of cases and deaths reported since 2014 with a number of seroprevalence studies around the country to inform mathematical models that estimate the underlying level of infection and cumulative levels of exposure in the population, the underlying patterns of disease and death by age and sex and the impact of vaccination^6^.

## Methods

### Data

We used an anonymized line list of all laboratory-confirmed chikungunya cases, confirmed via RT-PCR or serology, reported to the Laboratory Information System (*GAL - Gerenciador de Ambiente Laboratorial*) between January 2014 and September 2024 (N=488,234) (Table S1). We aggregated cases according to sex, age group, state, and year. We downloaded all death certificates citing chikungunya as a cause of death from the Mortality Information Service (*SIM - Sistema de Informação sobre Mortalidade*) over the same time period (N=1,719) ^7^. To help inform the probability that an infected individual seeks care, we collected data from 12 publicly available cross-sectional serological surveys conducted in Brazil between 2015 and 2023 (Figure 1A, Figure S1) ^8–19^. Finally, we downloaded sex- and age-stratified population estimates for each state from the Brazilian Institute of Geography and Statistics (*IBGE - Instituto Brasileiro de Geografia e Estatística*)^20^. We aggregated case, mortality, and population data by sex, age group, state, and year, using the age groups included in the case line list: <1, 1-4, 5-10, 11-15, 16-20, 21-25, 26-30, 31-35, 36-40, 41-45, 46-50, 51-55, 56-60, 61-65, 66-70, >70.

**Figure 1:**
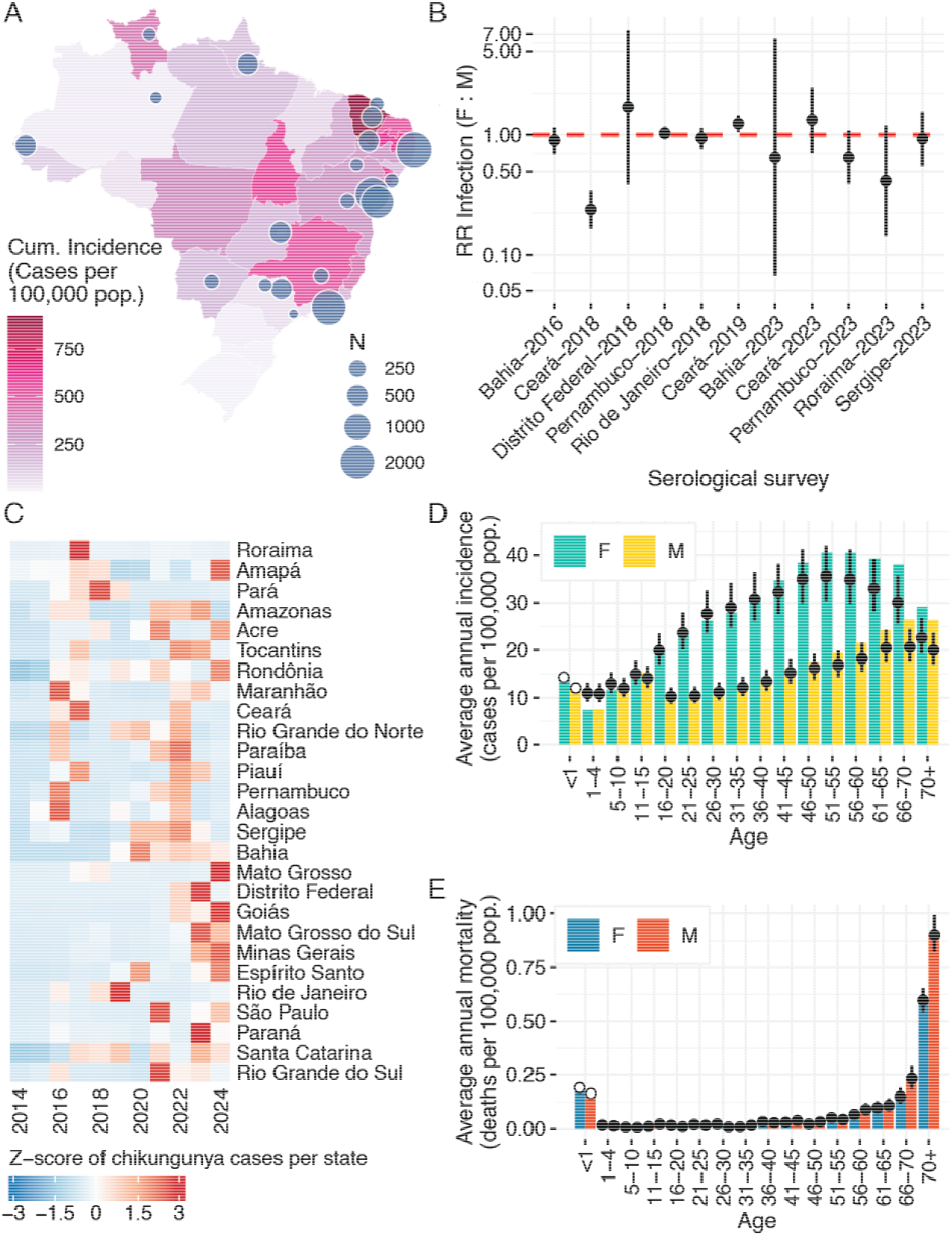
**(A)** Chikungunya incidence for all Brazilian states between 2014 and September 2024, with the location of publicly available CHIKV serological surveys. Point sizes indicate size of serological survey; **(B)** Relative risk (RR) of infection in females in comparison to males calculated from serological surveys with sex information. Values greater than one indicate higher risk of seroprevalence in females than in males in the corresponding study. Points represent median estimates and error bars represent 95% confidence intervals. **(C)** Z-scores of the number of chikungunya cases in each state across time. States are arranged according to their region and latitude, from North to South. **(D)** Average annual chikungunya incidence (in cases per 100,000 pop.) aggregated by age and sex in bars, with model fits represented by points (median estimates) and error bars (95% credible intervals). **(E)** Average annual chikungunya mortality (in deaths per 100,000 pop.) aggregated by age and sex in bars, with model fits represented by points (median estimates) and error bars (95% credible intervals). Chikungunya cases and deaths in <1 year olds were not included in our model and rates were estimated retrospectively.

### Model assumptions

We developed a mathematical model to estimate the annual force of infection (FOI) *A* for each of the 27 Brazilian states between 2014 and 2024. The FOI represents the rate at which individuals in the susceptible population get infected in a specific setting. We assumed equal risk of infection across sexes and age groups, which is consistent with the findings from serological surveys (Figure 1B, Figure S1).

Assuming that the population was entirely susceptible prior to 2014, the expected number of infections sex *s* and age group *a* in state *l*, in the year *t* is given by *λ*_*i,t*_ *S*_*s,a,l,t*_ *P*_*s,a,l,t*_, where *P*_*s,a,l,t*_, and *S*_, *s,a,l,t*_ represent the population and the proportion susceptible in that group. The expected number of reported cases 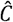 per cohort is then determined by multiplying the expected number of infections with the product of sex- and age-specific and of time-dependent probabilities of detection *ρ*_*s,a*_ x *ρ*_*t*_. We used the same age classes as in the data, and we assumed that, due to chikungunya becoming a notifiable disease in Brazil, probability of detection increased over time ^21^. In order to characterize sex- and age-specific probabilities of detection independently of time, we computed marginal probabilities of detection per sex *s* and age group *a*. Similarly, we computed marginal time-dependent probabilities of detection. Additionally, the expected number of deaths 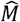 per sex *s* and age class *a* is determined by the accumulated number of infections in that group and age- and sex-specific infection fatality ratios *IFR*_*s,a*_.

### Likelihood and estimation

We fit our model with a joint likelihood where, for each serological survey *i*, with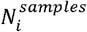 samples and 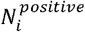 positive samples, 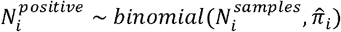, where 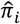 is the expected seroprevalence in the setting of each survey. Further, we assume that case counts *C* follow a negative binomial distribution with an estimated dispersion parameter 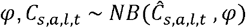, and that deaths *M* follow a Poisson likelihood 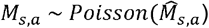 We fit the model in a Bayesian Markov Chain Monte Carlo (MCMC) framework using the cmdstanr package v2.36.0 in R v4.4.1. We generated four independent chains with 6,000 iterations each, discarding the first 1,000 iterations as warm-up. We considered that the model had converged if 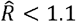 and if the effective sample size was greater than 300 when combining the posteriors of all chains.

### Sensitivity analyses

We conducted a sensitivity analysis to characterize spatial variations in the probability that suspected cases have confirmatory testing by incorporating suspected case counts from each state from the Notifiable Diseases Information System (Figure S2). Additionally, we conducted a sensitivity analysis where we fit the model using only laboratory-confirmed deaths reported in the Mortality Verification Service from the Brazilian Ministry of Health in order to provide IFR estimates based on serologically or virologically confirmed chikungunya deaths (Figure S3).

### Potential vaccine impact

The first chikungunya vaccine, IXCHIQ, was licensed in Brazil in April 2025. The vaccine was licensed through an accelerated pathway, and there are currently no estimates of vaccine efficacy from Phase IV clinical trials. We leveraged our model estimates to evaluate and compare the potential impact of different vaccine rollout strategies over the next five years. We defined a base case scenario of potential CHIKV circulation, vaccine, rollout campaign, and clinical chikungunya characteristics, and we conducted sensitivity analyses where we relaxed one parameter at a time to account for underlying uncertainty.

To capture the underlying infection risk, we simulated CHIKV circulation in each state by sampling FOI posterior estimates from the 2020-2024 time period for that state. We assumed that individuals who have been infected in the past have acquired life-long immunity ^22,23^. Though IXCHIQ has only been licensed for use in adults in Brazil, findings from safety and immunogenicity studies in adolescents have led to the extension of its license to individuals aged >12y in the European Union ^19,24^. Since the license in Brazil may be extended to adolescents in the future, in our base case scenario, we assumed that 40% of the population aged >12y was vaccinated at a national level between September and December of 2024, regardless of prior CHIKV infection status.

We assumed that the vaccine could confer life-long protection against infection and against disease. We informed all vaccine and clinical chikungunya parameters from previous modeling studies^1,25^. Simulating CHIKV circulation over five years with and without vaccination, we measured potential vaccine impact in terms of infections, cases, detected cases, chronic cases, deaths, and Disability-Adjusted Life Years (DALYs). In addition to our base case scenario, we evaluated the impact of vaccinating the population aged >50y at a national level and, for both age groups, we evaluated the potential impact of deploying the vaccine in the five states that have reported the highest chikungunya burden. Finally, in light of the US Food & Drug Administration recommending a pause in use of IXCHIQ in individuals aged >60y, we also conducted a sensitivity analysis targeting the national population aged 18-59y.

To account for considerable levels of uncertainty in future CHIKV circulation, in vaccine characteristics, and in the feasibility and reach of vaccination campaigns, we conducted sensitivity analyses where we incorporated variations in all simulation parameters, making them vary one at a time. Parameters used in the base case scenario and in sensitivity analyses are detailed in Table S3.

Further details of the model can be found in the Supplementary Materials.

## Results

We found considerable spatial heterogeneity in reported chikungunya case incidence across states since 2014 (Figure 1A). States in the Northeastern region reported the highest incidence, with 84 cases per 100,000 population per year in Ceará and 54 in both Paraíba and Sergipe. There were substantial differences in chikungunya incidence by sex, with 1.89 times more female confirmed cases than male cases, rising to 2.1 times more female cases in individuals aged >16y (Figure 1D). Chikungunya mortality was highest in infants and in older age groups with 0.18 deaths per 100,000 population per year in individuals aged <1y and 0.2 in individuals aged 65-70y, increasing to 0.72 in individuals aged >70y (Figure 1E). Mortality was higher in males than females (0.078 vs 0.071 per 100,000 per year).

Across the 12 serological surveys identified in our study, we found no difference in the underlying levels of infection by age or sex. Of the 11 settings with sex-stratified information, nine had 95% confidence intervals of the relative risk (RR) of infection by sex that included 1, suggesting no significant differences (Figure 1B). Among the remaining two settings, one found a higher RR of infection in males, and the other one found a higher RR in females.

Using the case incidence data, mortality, and seroprevalence data we fitted a mathematical model to estimate the annual FOI by year and state, as well as probabilities of infections being detected by age and sex and by time. We found the mean FOI was 2.1% per year (95%CrI: 1.7-2.6) at a national level, with an estimated 40.9 (95%CrI: 33.2-50.6) million infections having occurred between January 2014 and September 2024, representing 18.3% (95%CrI: 16.5-20.3) of the Brazilian population. There was substantial variation in mean FOI values across states (Figures 2C, S4A). Ceará and Paraíba had the highest mean FOI, with an average of 8.8% (95%CrI: 7.6-10.2) and 8.7% (95%CrI: 6.6-11.4) of the susceptible population getting infected annually. Given the sporadicity and variation in magnitude of CHIKV outbreaks, most of the burden in each state was accumulated in 1-5 of the 11 years since 2014 (Figure S4B). However, we found no evidence of a subsequent reduction in CHIKV FOI in states with high levels of accumulated infection in the population (Figure S5).

**Figure 2:**
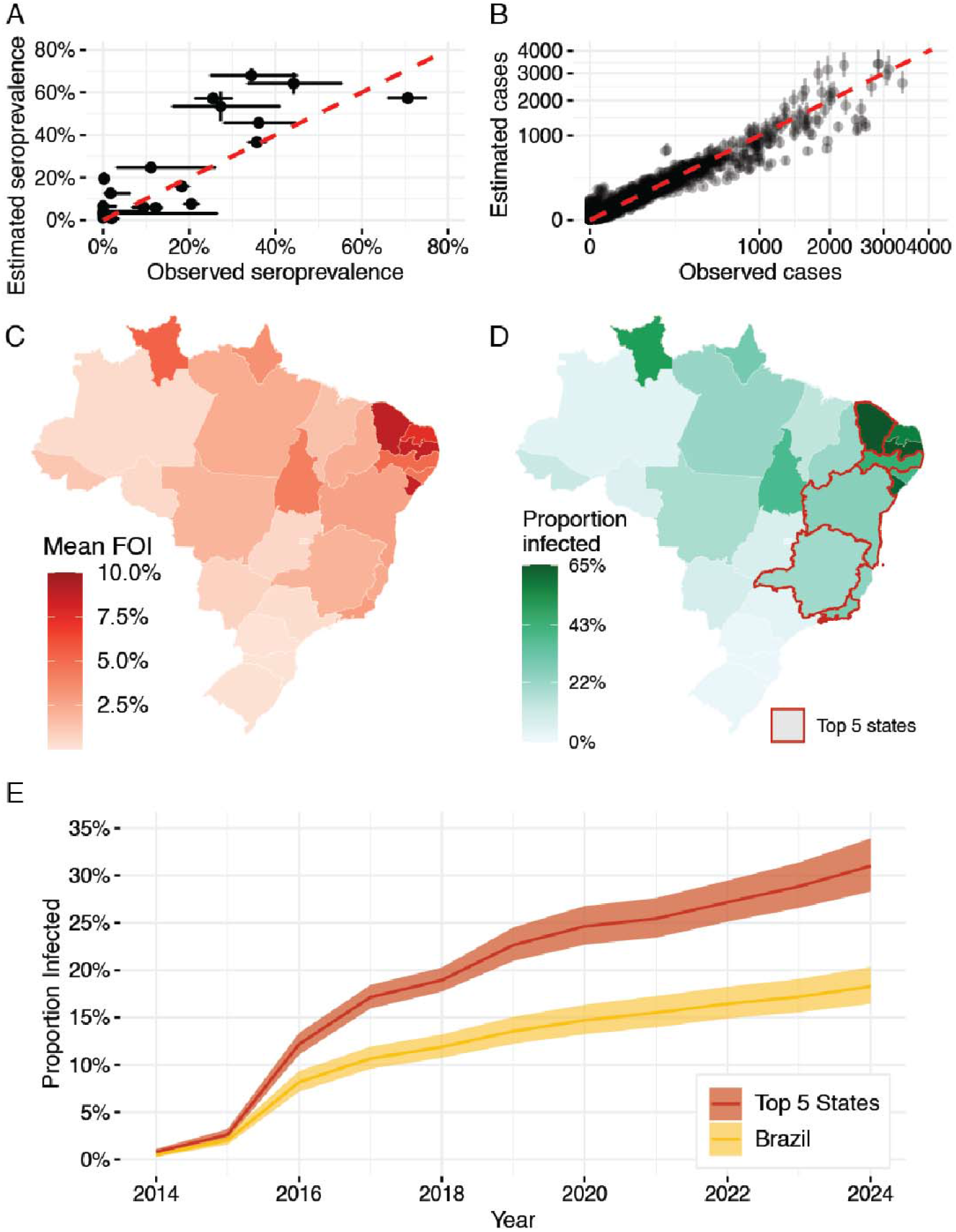
**(A)** Estimated seroprevalence in the setting of each serological survey included in our model as a function of the seroprevalence observed in each survey; **(B)** Estimated number of cases for each case data point included in our model as a function of observed number of cases; **(C)** Mean CHIKV attack rate per Brazilian state between 2014 and 2024; **(D)** Estimated level of population infected with CHIKV in each state at the end of September 2024. The top five states in terms of number of cases reported are highlighted: Minas Gerais, Ceará, Bahia, Rio de Janeiro, and Pernambuco. **(E)** Average proportion of the population infected as a function of time for Brazil and for the top five states. Lines represent median estimates and ribbons represent 95% credible intervals.

Overall, we estimated that 1.13% (95%CrI: 1.07-1.19) of infections were detected by surveillance systems, ranging from 0.024% (95%CrI: 0.019-0.03) in 2014 to 3.6% (95%CrI: 2.9-4.4) in 2024 (Figure 3A). On average, 1.45% (95%CrI: 1.38-1.53) of female infections were detected compared to 0.79% (95%CrI: 0.75-0.84) of male infections. The probabilities of detection were comparable in male and female infections between the ages of 1 and 15y (Figure 3D). The marginal probability of detection in females followed a log-linear trend from ages 1-4y to ages 51-55y, with every five-year increase in age representing a 14.3% increase in the probability of detection (Figure S6). Contrastingly, the probability of detecting male infections dropped after the age of 15y from 0.76% to 0.55% before following a log-linear increase from ages 16-20y to 61-65y. Each five-year increase in age was associated with a 10.1% increase in the probability of detecting infections There was a significant difference in detection probability between the sexes across all age groups over 16y. The highest difference appeared in individuals 26-30y, with a RR of 2.5 (95%CrI: 2.2-2.7). Differences declined after ages 51-55y, reaching almost comparable levels in individuals aged >70y (RR 1.15, 95%CrI: 1.05-1.27).

**Figure 3:**
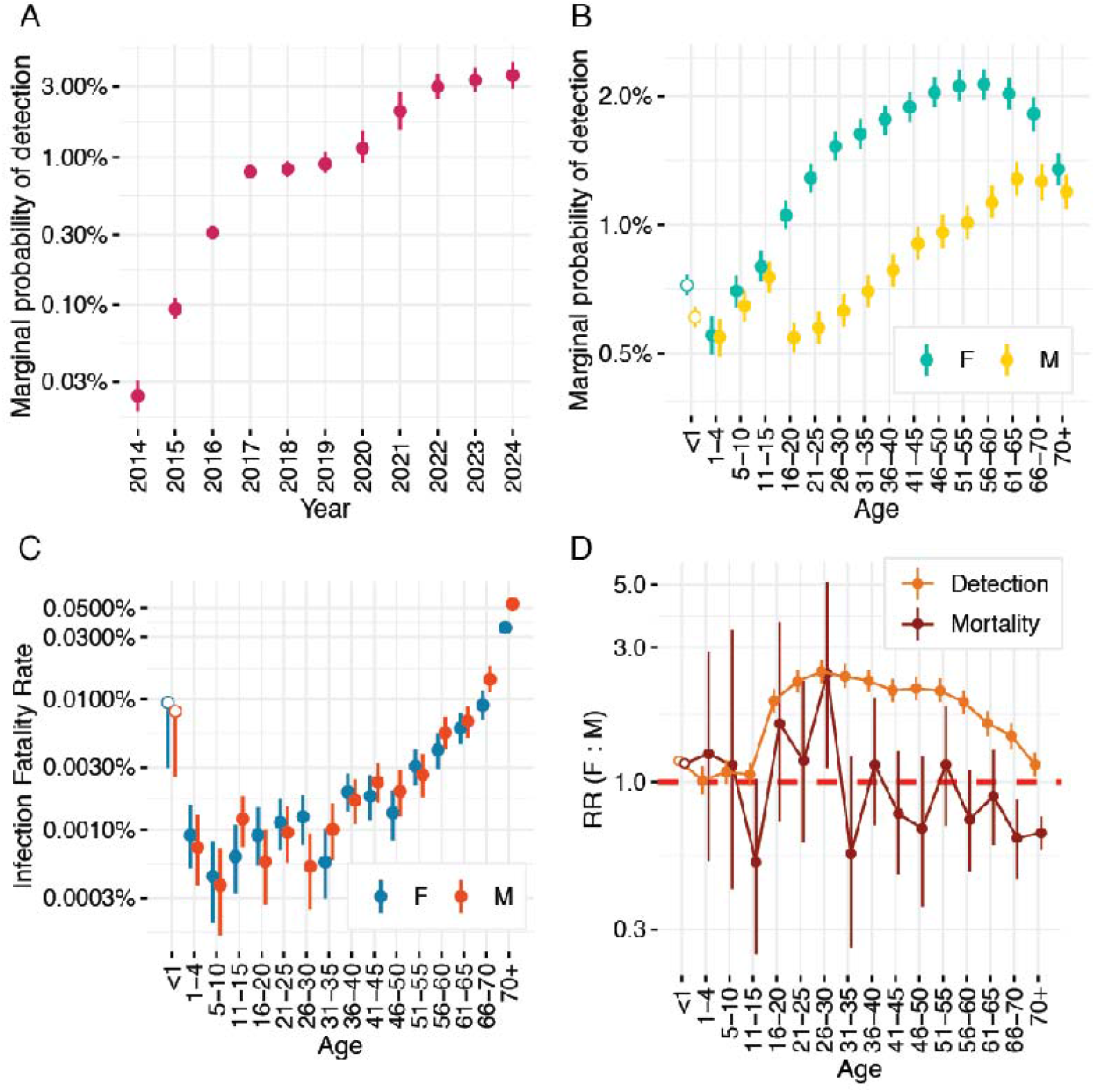
**(A)** Marginal time-dependent probability of detection given infection; **(B)** Marginal sex- and age-dependent probability of detection given detection; **(C)** Age- and sex-dependent IFR; **(D)** Relative risk (RR) of detection and mortality in females in comparison to males as a function of age. Probabilities of detection, IFRs, and RRs are plotted in the log scale. Probabilities of detection, IFRs, and RRs in <1yo were estimated retrospectively. Points represent median estimates and error bars represent 95% credible intervals.

On average, 0.0042% (95%CrI: 0.0039-0.0045) of CHIKV infections resulted in death. IFR estimates were highest in infants, at 0.0087% (95%CrI: 0.0029-0.0094), and in older age groups, with 0.011% (95%CrI: 0.009-0.014) in individuals aged 66-70y, and 0.043% (95%CrI: 0.040-0.047) in individuals aged >70y. From the age of 50y onwards, each year increase in age was associated with a 10.7% (95% CI 8-13.4) increase in the probability of death given CHIKV infection. IFR estimates were similar across female and male infections for most age groups, with differences appearing in older age groups (Figures 3C-D). In individuals aged >66y, the risk of death given infection in males was 1.5 times (95%CI: 1.2-1.9) the risk of death in females. A sensitivity analysis using only laboratory confirmed chikungunya deaths resulted in a mean IFR of 0.0021% (95%CI: 0.0019-0.0025) (Figure S3).

Our estimates rely on laboratory-confirmed cases that have been reported by public health laboratories nationally. We quantified spatial heterogeneities in laboratory ascertainment through a separate model where we incorporated state-level aggregates of suspected cases across time (Figure S2). We found considerable variation in the probability that a suspected case will ultimately be confirmed through laboratory testing, with values ranging from 13% in the Federal District of Brasilia, to 62% in Acre.

Using our model estimates of the underlying probability of infection and the probability of symptoms and death following infection, we investigated the potential impact of a CHIKV vaccine over five years, exploring rollout strategies where either the populations aged >12y or >50y would be targeted, regardless of prior CHIKV infection status, both at a national level or in the five states that have accumulated the most cases (Figure 4). We found that to vaccinate 40% of the target population with a single-dose vaccine ranged from 8.4 million doses when targeting individuals aged >50y in the top five states to 73 million doses for a nationwide vaccine campaign in individuals aged >12y. Assuming that transmission between 2020 and 2024 is representative of CHIKV circulation over the next five years, and that a vaccine would provide lifelong protection against infection of 70% and against symptomatic disease of 95%, we estimated that targeting the population aged >12y at a national level would be the most impactful strategy in terms of burden averted, averting 2.3 million (95%CrI: 0.7-4.2 million) infections, 1.6 million (95%CrI: 0.5-3.0 million) cases, 77,000 (95%CrI: 23,900-142,000) chronic cases, and 197 (95%CrI: 61 - 359) deaths, amounting to 21,900 (95%CrI: 6,820-40,400) DALYs averted. However, measuring impact in terms of burden averted per 10,000 vaccine doses used would favor targeting the population in the top five states. We estimated that 380 (95%CrI: 123-681) cases would be averted per 10,000 doses should the population aged >12y be targeted in these states, compared to 371 (95%CrI: 118-665) if the vaccine were rolled out to the population over the age of 50y. The latter strategy would have a higher impact in terms of deaths averted per 10,000 doses (0.12 [95%CrI: 0.039-0.21] for the >50y strategy vs 0.048 [95%CrI: 0.016-0.084] for the >12 strategy).

**Figure 4:**
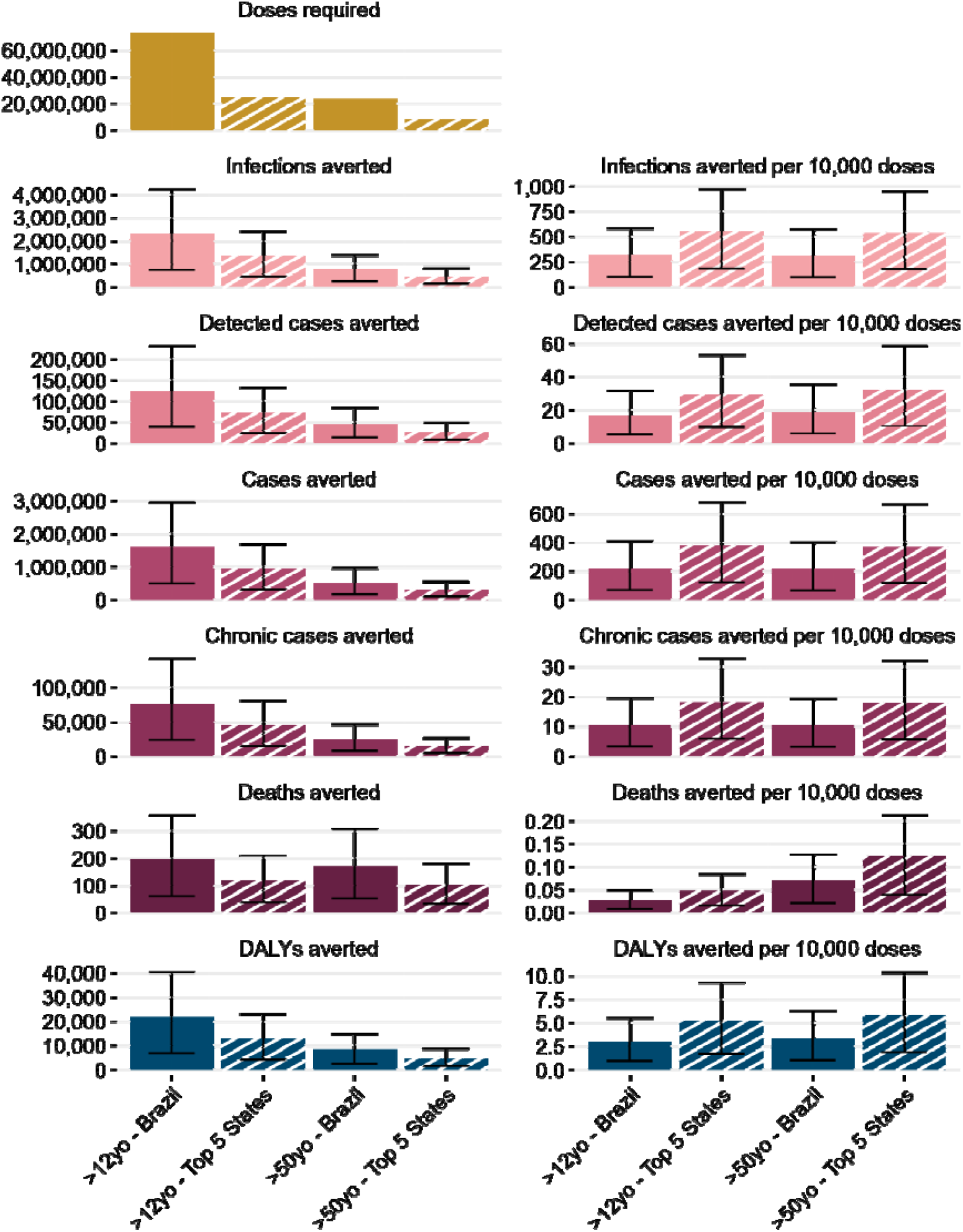
Doses used and potential averted burden in terms of infections, detected cases, all cases, chronic cases, deaths, and DALYs for different vaccine rollout strategies over the next five years. Bars represent median estimates and error bars represent 95% credible intervals. Averted burden is also presented as burden averted per 10,000 vaccine doses used over the five-year period.

We conducted sensitivity analyses where we introduced variations in each parameter of our vaccine impact simulations (Figure 5, Table S3). Using vaccination in the population aged >12y at a national level as our base case scenario, we compared the number of cases, deaths, and DALYs averted in each simulation. Targeting individuals aged 18-59y, which would align with FDA recommendations at the time of writing, would result in a 28% reduction in cases averted and of 77% of deaths averted compared to the base case scenario. We find that our projections of vaccine impact are most sensitive to assumptions around duration of protection, vaccine efficacy, vaccine coverage, target population, and around the probability of developing symptomatic disease and of death following infection.

**Figure 5:**
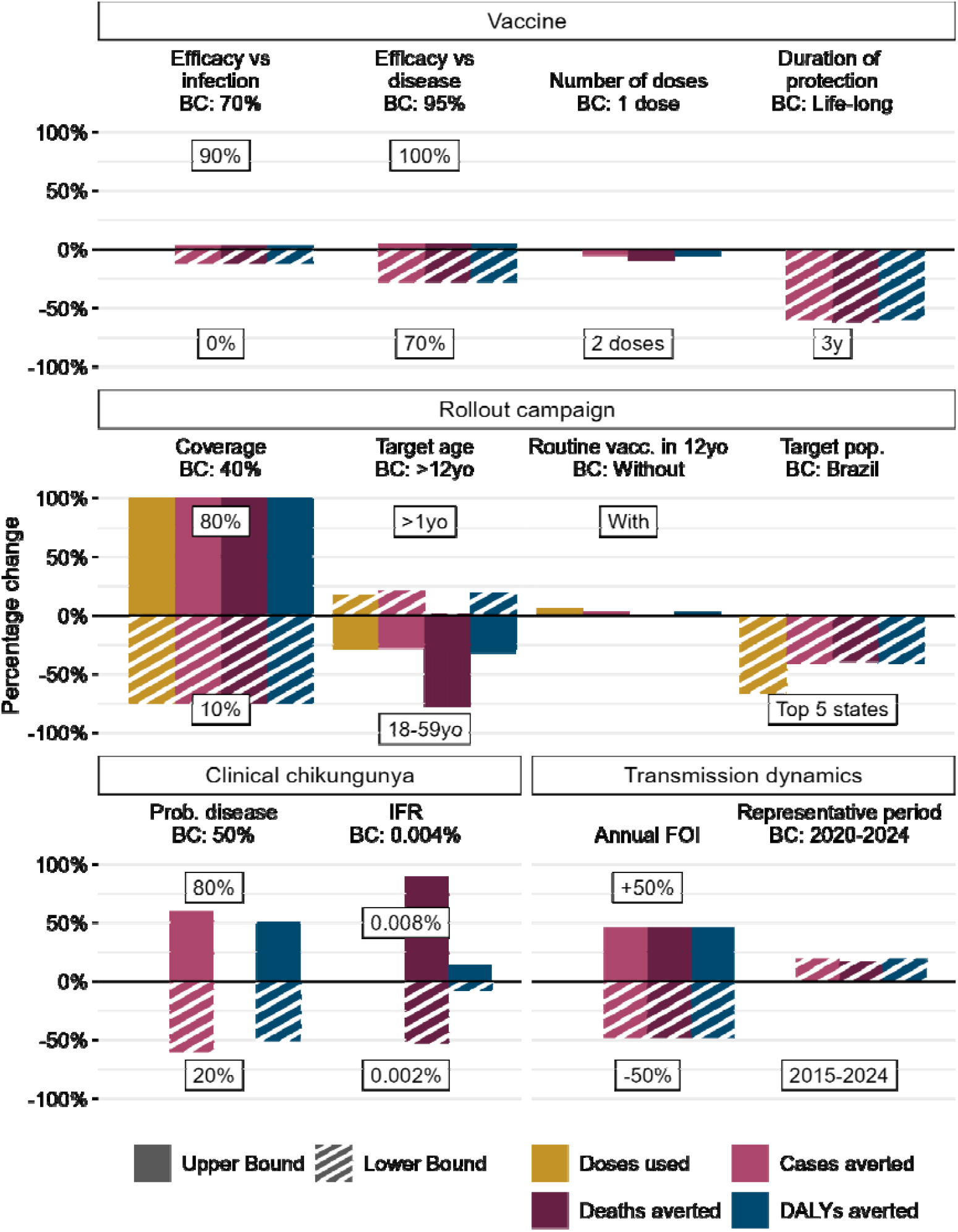
Sensitivity analysis of differences in potential vaccine impact when one parameter changes in comparison to the base case (BC) scenario. Comparisons are represented as the percentage change of median estimates of doses used, and cases, deaths, and DALYs averted.

## Discussion

Here we combined age-specific case data, death data, and seroprevalence data within the same analytical framework to reconstruct CHIKV transmission across Brazil as it became endemic. We estimate that over 40 million people in the country have been infected with CHIKV, representing 18% of the population, especially in the northeastern region. Introduction of a vaccine could substantially reduce the burden from the pathogen, especially if targeted to the areas with a history of high CHIKV circulation. We found that vaccinating individuals aged >50y is the most effective use of the vaccine. However, the risk in younger individuals remains substantial, especially of potentially chronic, debilitating arthralgia^26^. Therefore, focusing on wider age groups may remain a priority. The likely exclusion of individuals >60y from vaccine programs would limit the potential impact of the vaccine, as nearly all chikungunya related deaths are in this oldest age group.

Our estimates of the average FOI are consistent with global estimates in endemic locations, (estimated at 2.4% compared to 2.1% here)^1^. Our study also identified divergent patterns of disease risk by age and sex, despite no differences in underlying infection risk, consistent with fundamental differences in the probability of developing severe symptoms. Underlying differences between males and females, especially after puberty until reproductive senescence, have been observed for a number of pathogens ^29^. These patterns match hormonally driven differences in immune responses, which mean women are more susceptible to inflammatory disease, including arthralgia ^30^. While we cannot wholly exclude a role of differences in health-seeking behavior, previous outbreak investigations, including studies with systematic household surveys, report similar differences in sex- and age-dependent disease probabilities ^31–33^. Higher mortality in males in older age groups could be driven by faster immunosenescence or, alternatively, immunosenescence could make women less vulnerable by reducing an initially stronger immune response. These sex- and age-dependent patterns of disease and death risks exemplify the male-female health survival paradox, but the interplay between hormonal and chromosomal differences across sexes and ages has not been studied in CHIKV ^34^.

Mortality was also highest in infants and the elderly, consistent with classical ‘U’-shaped risks of death by age. A remarkably consistent 10% annual increase in the probability of death following infection has also been found for other pathogens, including coronaviruses (SARS-CoV-2, MERS) and influenza ^29^, and now appears to be relevant for CHIKV too. The reasons for this log-linear increase by age are unclear but may relate to immune function, including a reduced ability to clear infections. In general, we estimate that around 4 in 100,000 infected individuals die, confirming CHIKV’s deadly nature ^35,36^. Our estimates are lower than those from a recent outbreak in Paraguay, where a national seroprevalence study conducted immediately after the outbreak found around 13 deaths per 100,000 individuals infected^37^. It is unlikely that underlying mortality risk differs significantly between two neighbouring countries with the same circulating strains. Our findings suggest that there may be ‘missing’ CHIKV deaths in Brazil attributed to other causes. Consistent with this hypothesis, is the finding of excess mortality during high chikungunya years^38^. In addition, COVID-19 deaths were found to be consistently under-reported across the Americas in the elderly^39^. Alternatively, our FOI estimates may be too high. We note that we overestimate seroprevalence in some settings, which impacts calibration of the underlying level of infection from observed case numbers. We made a necessary assumption that seroprevalence estimates from a location are representative of serostatus across the whole state. However, we acknowledge there are substantial differences in infection risk across neighbouring communities ^5^.

While studies such as this one improve our understanding of underlying infection risk and of the risks of acute disease and death from infection, understanding of arthralgia risk remains poor. Here we rely on estimates from literature reviews, which draw primarily from cohorts of clinically attended chikungunya^25^. However, the generalizability of these estimates to Brazil is unclear. A key barrier is the lack of surveillance datasets recording CHIKV-associated arthralgia. By the time individuals seek care for long term arthralgia, CHIKV infection is often cleared, and symptoms are not sufficiently specific to allow diagnosis. Systematic follow-up of confirmed acute infections to quantify the incidence of subsequent arthralgia will help improve these estimates, which could then be integrated into models. Further, there is a growing understanding of central nervous system (CNS) infections as a potential severe consequence of CHIKV infection ^45,46^. Specific recording of CNS disease from CHIKV infection will allow us to consider this severe outcome in quantifying the potential vaccine benefit.

Our study is subject to limitations. Both currently approved vaccines, IXCHIQ and VIMKUNYA, were approved through neutralizing antibodies correlated with protection^40^. Importantly, these correlates are based on human cohorts and non-human primate passive transfer studies ^41–44^. Without traditional Phase III efficacy trials, we lack vaccine efficacy estimates to guide our models. Instead, we rely on expert opinion to guide likely efficacy and duration parameters^1^. Planned phase IV trials will be critical to obtain effectiveness estimates and further improve models. Further, we did not consider the risk of vaccine associated severe adverse events. The first mass use of IXCHIQ was during an outbreak in La Reunion in 2024/25, where over 51,000 cases and at least 12 deaths were reported^47^. The vaccine campaign was, however, suspended in individuals aged >65y following the deaths of three elderly vaccine recipients, although only one of the deaths has currently been linked to the vaccine^48^. A good understanding of the underlying risks from a vaccine is critical to its appropriate usage ^49^. A further limitation is that we assumed that the probability that a suspected CHIKV infected individual sought care was constant across states (but that it could vary by age, sex and year). In addition, some suspected cases will not have been from CHIKV, but potentially from dengue virus or from another cause ^50^. Similarly, some suspected dengue cases will have been instead caused by chikungunya. A substantial level of misdiagnosis would impact our estimates of the probability of a CHIKV infected individual seeking care. We note wide variability in the probability of case confirmation by PCR or ELISA across states, meaning we could not rely on confirmed cases only.

Despite these limitations, this study provides a robust evidence base of the underlying epidemiology of CHIKV since its introduction and for the development of a vaccine investment case for CHIKV vaccines in the country. It highlights that despite widespread transmission, most of the country remains susceptible, and that vaccination, especially in the Northeast and Southeast of the country, could limit the future burden.

## Data Availability

All data produced in the present study are available upon reasonable request to the authors.

## Ethics and data/code statements

This project was developed using publicly available data and therefore did not require ethical approval. Data and code to reproduce our analyses can be found in the following GitHub repository: [Will be provided upon acceptance].

## Author contributions

OCA conducted the formal analysis. OCA, MOD, and HS conducted the investigation and developed methods. STSdL, RdJ, and OCA collected and curated the data. OCA and HS wrote the initial draft, and MOD, GRdS, RdJ, STSdL, DS, CM, AD, MUG, and WMdS reviewed and edited the manuscript. HS supervised the project.

## Acknowledgements

This work was funded by the Coalition for Epidemic Preparedness Innovations (CEPI)

## Supplementary Figures

**Figure S1:**
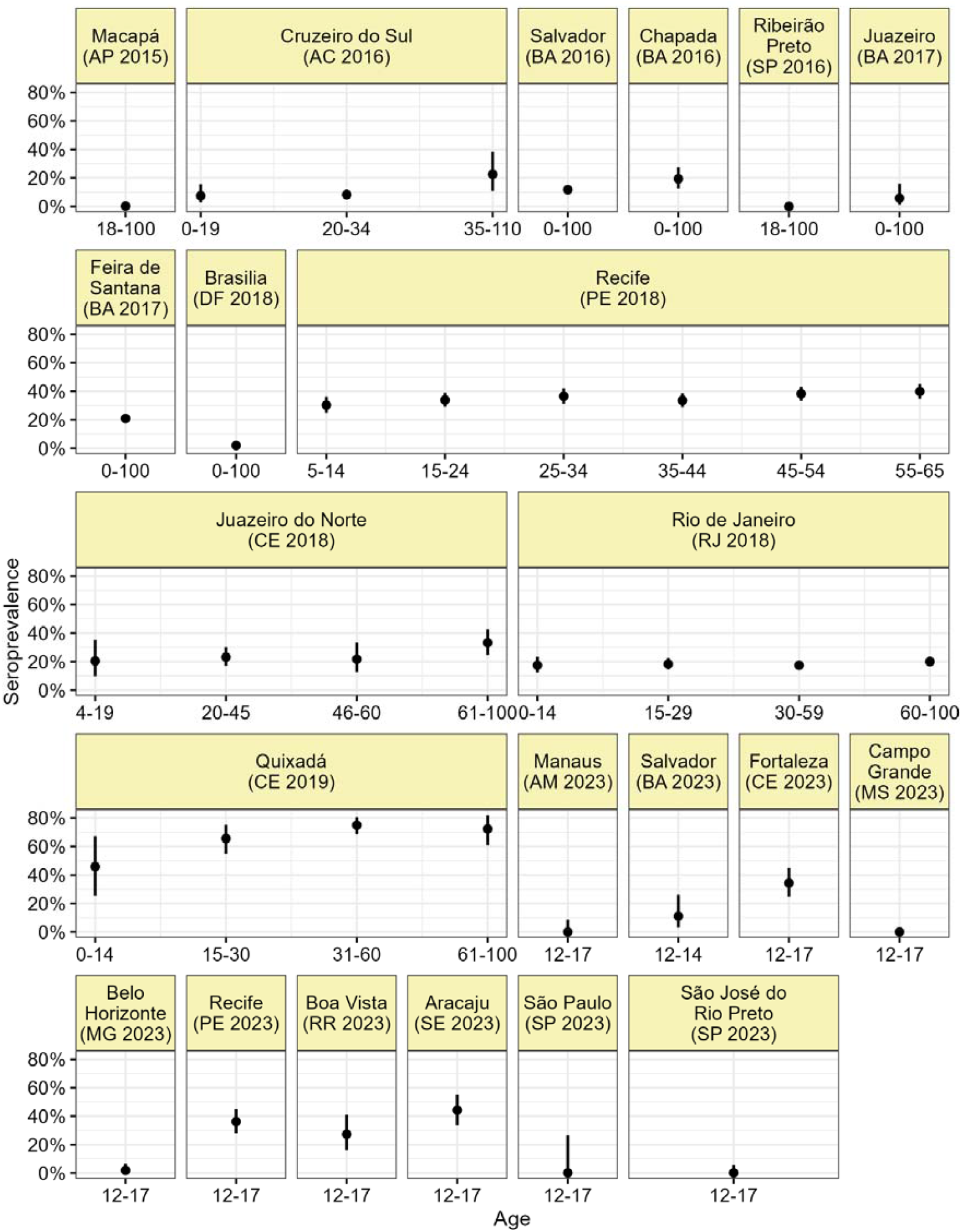
Seroprevalence found by publicly available serological surveys as a function of age.

**Figure S2:**
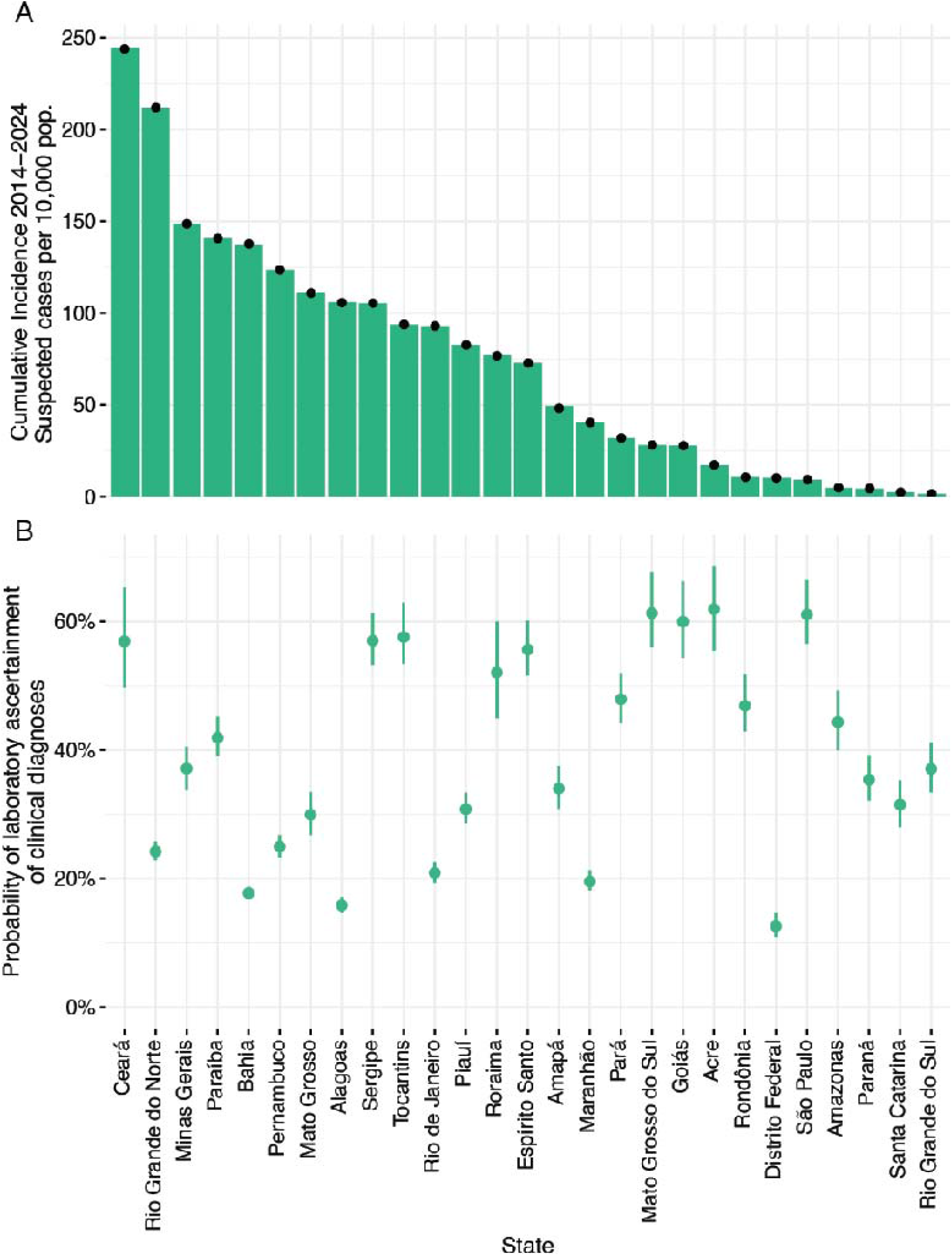
**(A)** Incidence of suspected cases per 10,000 inhabitants in each Brazilian state. Bars represent data and points and error bars represent median model estimates with 95% credible intervals. **(B)** Estimated probability of laboratory confirmation for suspected cases in each state. Points represent median model estimates and error bars represent 95% credible intervals.

**Figure S3:**
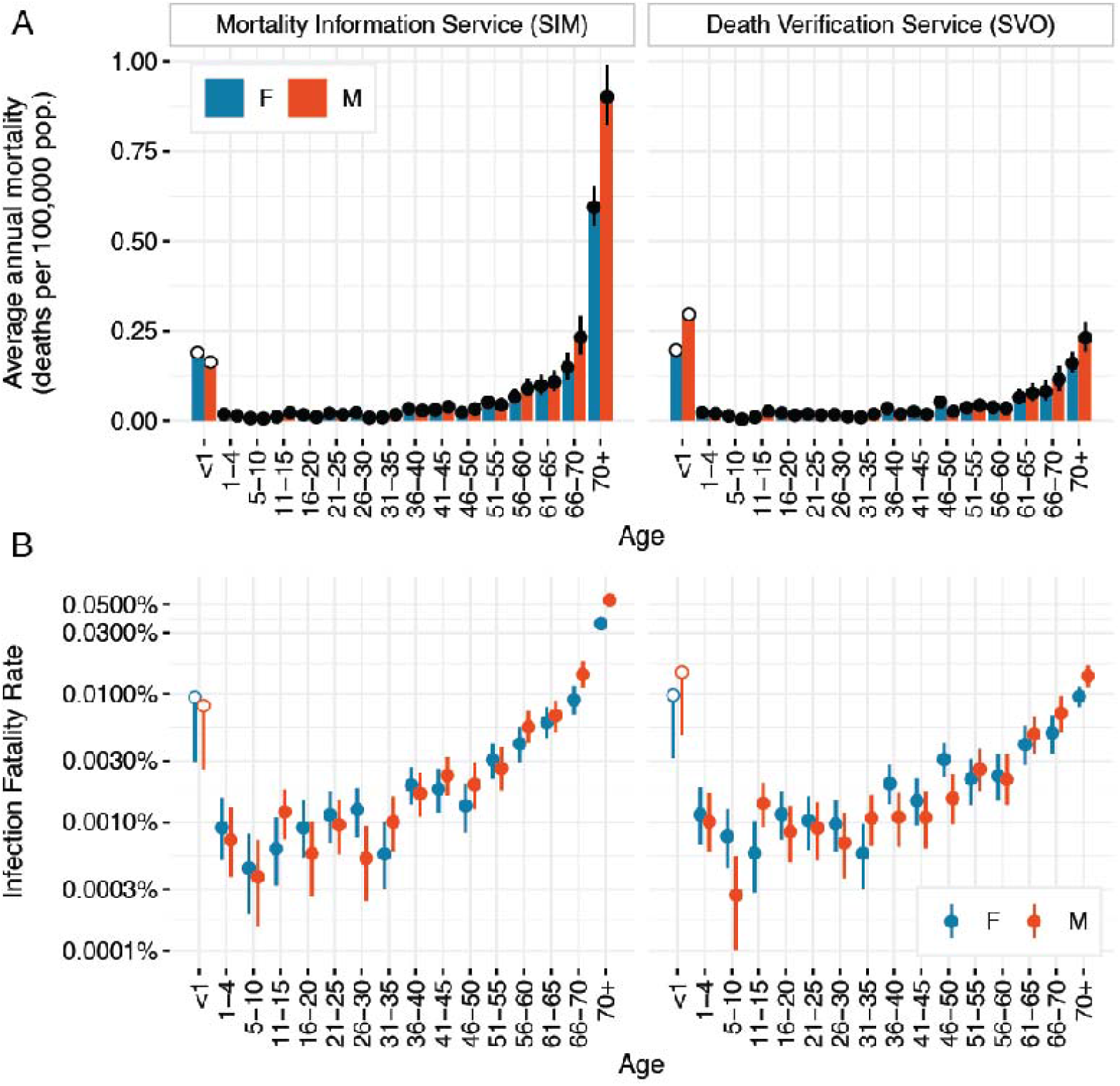
Comparison of sex- and age-dependent **(A)** mortality (in average annual deaths per 100,000 pop.) with model fit and **(B)** IFR when using chikungunya deaths reported in death certificates on the Mortality Information Service versus using laboratory confirmed deaths reported on the Death Verification Service. Points represent median estimates and error bars represent 95% credible intervals. Deaths in <1 year olds were not included in the model and estimates were evaluated retrospectively.

**Figure S4:**
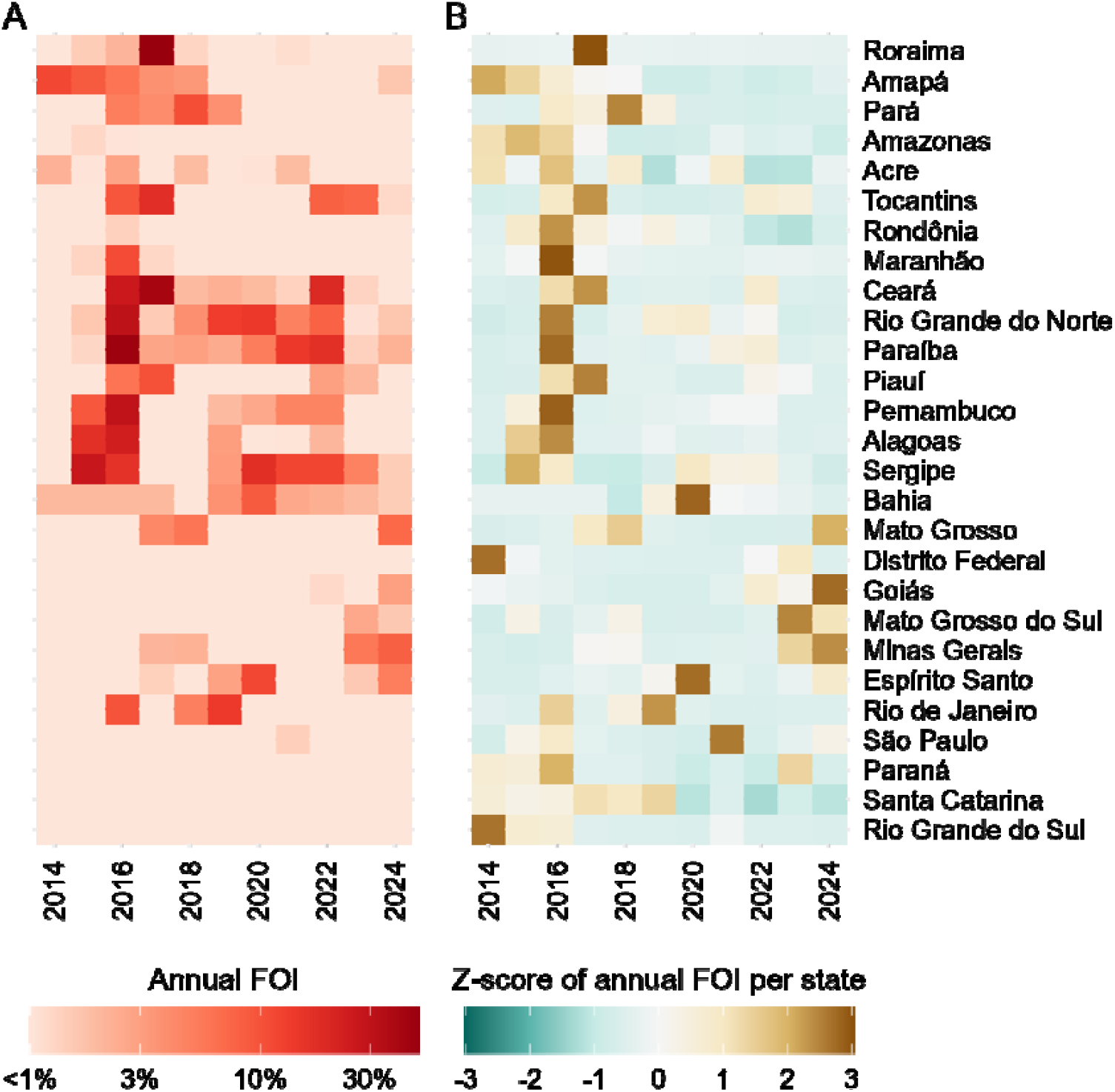
**(A)** Median estimate and **(B)** z-score of annual CHIKV FOI per state across time. States are arranged according to their region and latitude, from North to South.

**Figure S5:**
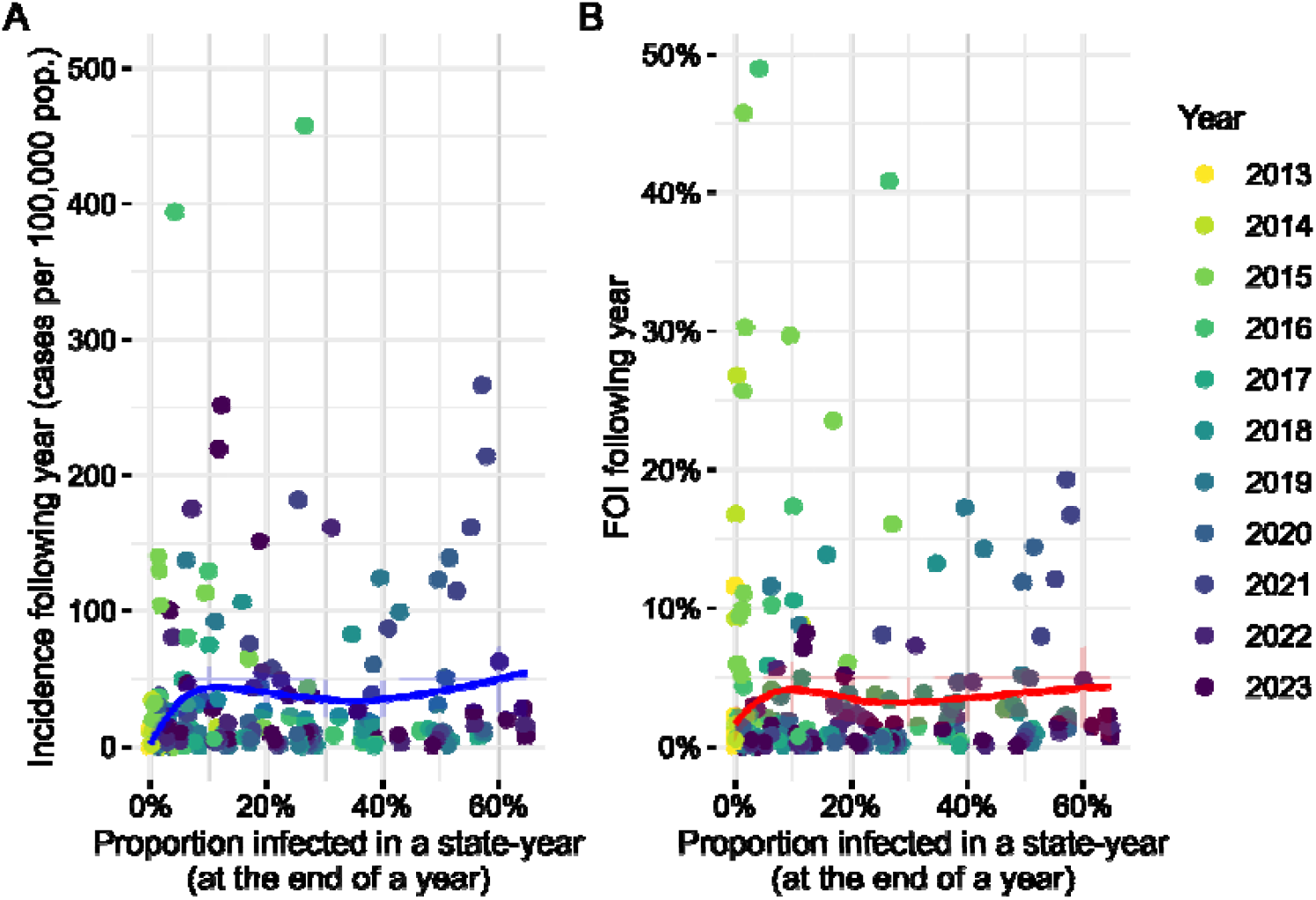
**(A)** Observed incidence (in cases per 100,000 pop.) and **(B)** estimated annual CHIKV FOI in a state as a function of the estimated proportion of the population that has been infected with CHIKV in the corresponding state at the end of the previous year. Lines represent model prediction and ribbons represent 95% confidence intervals of a LOESS regression.

**Figure S6:**
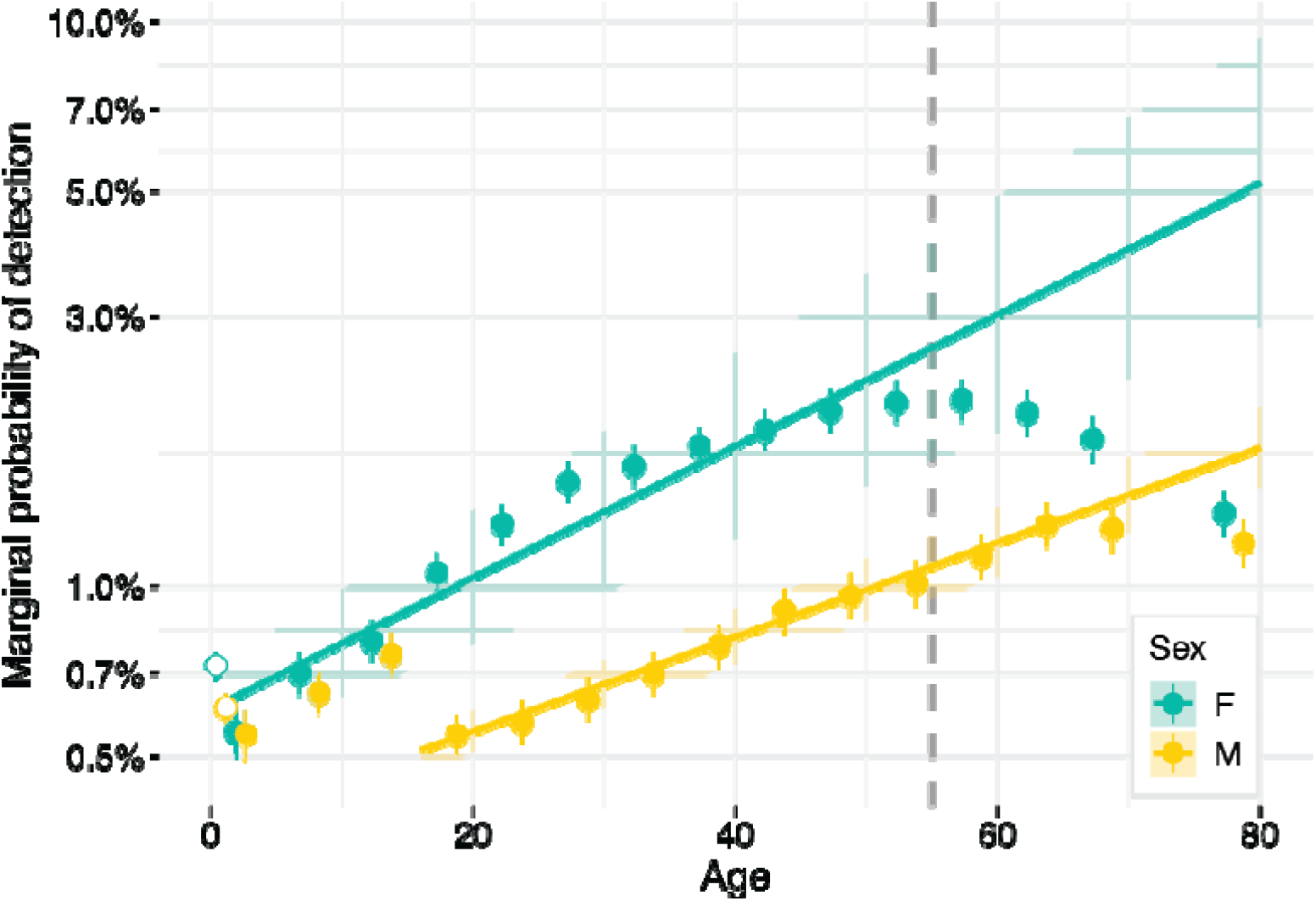
Log-linear regressions of the marginal probabilities of detection according to sex and age. Points represent median estimates from our model, and error bars represent 95% credible intervals. Lines and ribbons represent predictions and 95% confidence intervals from log-linear models. Log-linear regressions were fit using reporting probabilities from ages 1-4 in female infections, and from ages 16-20 in male infections, until ages 51-55 for both sexes (vertical gray line).

**Table S1:** Cases, deaths, marginal probability of reporting, and IFR for different demographic groups. Reporting probabilities and IFRs are reported with 95% credible intervals.

| Group | Cases | Deaths | Reporting probability (%) | IFR (%) |
| --- | --- | --- | --- | --- |
| SEX |  |  |  |  |
| Female | 319,117 | 837 | 1.45 (1.38 - 1.53) | 0.00393 (0.00363 - 0.00428) |
| Male | 169,117 | 882 | 0.791 (0.748 - 0.835) | 0.00439 (0.00493 - 0.00479) |
| AGE |  |  |  |  |
| <1 | 4,237 | 57 | 0.647 (0.212 - 0.695) | 0.00871 (0.00285 - 0.00935) |
| 1-4 | 9,617 | 20 | 0.549 (0.506 - 0.594) | 0.000832 (0.000545 - 0.00127) |
| 5-10 | 22,861 | 13 | 0.674 (0.627 - 0.718) | 0.000415 (0.000242 - 0.000653) |
| 11-15 | 24,926 | 28 | 0.777 (0.725 - 0.835) | 0.000928 (0.000619 - 0.00131) |
| 16-20 | 27,005 | 24 | 0.795 (0.746 - 0.854) | 0.000740 (0.000490 - 0.00107) |
| 21-25 | 32,276 | 35 | 0.931 (0.867 - 1.00) | 0.00106 (0.000742 - 0.00144) |
| 26-30 | 35,472 | 29 | 1.09 (1.01 - 1.16) | 0.000903 (0.000609 - 0.00126) |
| 31-35 | 37,915 | 25 | 1.17 (1.10 - 1.27) | 0.000795 (0.000530 - 0.00113) |
| 36-40 | 41,183 | 56 | 1.29 (1.20 - 1.38) | 0.00184 (0.00137 - 0.00235) |
| 41-45 | 42,010 | 57 | 1.42 (1.32 - 1.52) | 0.00206 (0.00158 - 0.00265) |
| 46-50 | 41,611 | 40 | 1.53 (1.42 - 1.64) | 0.00166 (0.00122 - 0.00221) |
| 51-55 | 40,655 | 64 | 1.59 (1.48 - 1.71) | 0.00288 (0.00227 - 0.00364) |
| 56-60 | 36,801 | 90 | 1.67 (1.55 - 1.79) | 0.00478 (0.00394 - 0.00586) |
| 61-65 | 30,884 | 98 | 1.69 (1.56 - 1.82) | 0.00634 (0.00515 - 0.00779) |
| 65-70 | 24,243 | 138 | 1.57 (1.46 - 1.69) | 0.0114 (0.009402 - 0.0136) |
| >70 | 36,538 | 945 | 1.29 (1.19 - 1.39) | 0.0430 (0.0397 - 0.0467) |

**Table S2:** Priors used in the mathematical model.

| Parameter | Scale | Prior |
| --- | --- | --- |
| Force of Infection ($\lambda$) | Log | Normal(-6, 1) |
| Probability of developing and reporting symptoms ($\rho$) | Logit | Normal(-5, 0.5) |
| Probability of laboratory confirmation given reported disease ($\psi$) | Linear | Beta(1.5, 1.5) |
| Infection fatality rate (IFR) | Log | Normal(-8,1) |

**Table S3:** Parameters used in vaccine impact simulations, with ranges used in sensitivity analysis

| Parameter | Base Case | Sensitivity |
| --- | --- | --- |
| VACCINE CHARACTERISTICS |  |  |
| Number of doses | 1 | 2 |
| Duration of protection | Life-long | 3 year half-life |
| Efficacy against infection | 70% | 0% - 90% |
| Efficacy against disease | 95% | 70% - 100% |
| ROLLOUT CAMPAIGN |  |  |
| Coverage | 40% | 10% - 80% |
| Target age | >12yo | >1yo - >50yo |
| Target population | Brazil | Top five states |
| Annual vaccination in 12yo | Excluded | Included |
| CLINICAL CHIKUNGUNYA |  |  |
| Probability of developing disease | 50% | 20% - 80% |
| IFR | 0.004% | 0.002% - 0.008% |
| TRANSMISSION DYNAMICS |  |  |
| Amplification of annual FOI | None | -50% - +50% |
| Representative period | 2020 - 2024 | 2015 - 2024 |

## Supplementary Methods

### Data

We excluded cases from the GAL line list and death certificates from the SIM database for which at least one of the relevant variables (sex, age group, municipality, date of symptom onset, laboratory ascertainment technique) had not been recorded, as well as those that had been confirmed by IgG serology, as this type of antibody is less indicative of acute infection.

The serological surveys included in our analysis were identified through a literature review detailed in a previous study ^1^.

### Model assumptions

We used the seroprevalence studies, with data from 13 states, assuming that seroprevalence found in the municipality where each serological survey was conducted was representative of the state-wide seroprevalence in that given year. Since there were no significant differences in seroprevalence across different age groups in included surveys (Figure S1), we aggregated the results of different age groups to one single seroprevalence observation per study. Thus, for each survey *i*, with seroprevalence observations for age groups in *A*_*i*_, the total number of samples 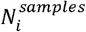 and the total number of positive samples 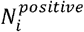 are determined by:

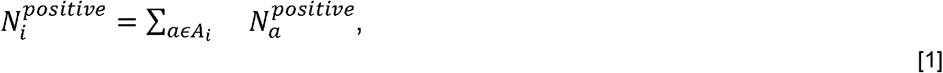

And

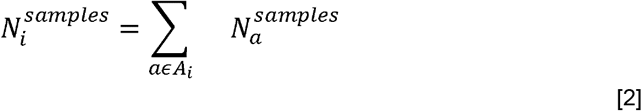

Assuming that the population was entirely susceptible prior to 2014, the expected number of cases 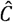 of sex *s* and age group *a* in state *l*, in the year *t* is given by

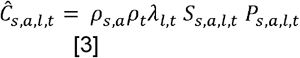

where *P*_*s,a,l,t*_ and *S*_*s,a,l,t*_ represent the population and the proportion susceptible in that group. We additionally infer age- and sex-specific infection fatality ratios (IFR) by fitting to the cumulative number of CHIKV deaths by age and sex reported in Brazil. The expected number of deaths 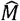 per sex and age group is given by:

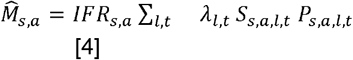

In order to characterize sex- and age-specific probabilities of detection independently of time, we computed marginal probabilities of detection per sex *s* and age group *a* as follows:

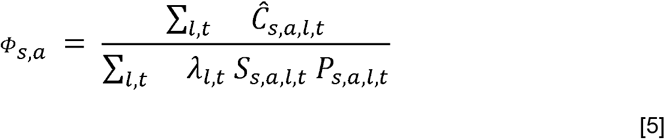

Similarly, we computed marginal time-dependent probabilities of detection independently of sex and age per year t:

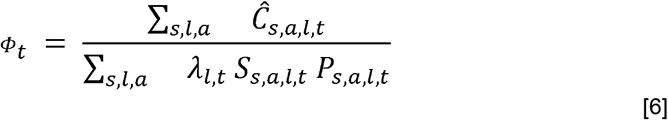

Moreover, let {*σ*_*i*_, *A*_*i*_} denote the sexes and age groups from which samples were collected in survey *i*, and *L*_*i*_ and *T*_*i*_ denote the state and the year in which the serological survey was conducted. Using these, we compute the expected seroprevalence 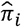 according to the setting of the study:

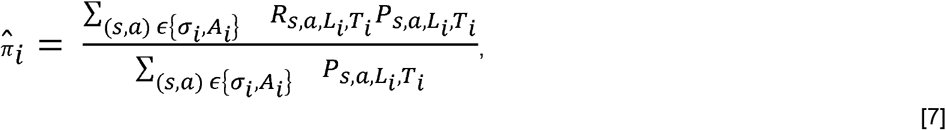

where *R*_*s,a,l,t*_ denotes the proportion of the population of sex *s* and age group *a*, in state *l*, that has been infected by year *t*.

### Likelihood and estimation

In the first four years of endemic CHIKV circulation (2014-2017) negligible cases were reported in the state of Bahia (N=135) despite seroprevalence studies indicating 12.2% (95% CI 10.8-13.8) and 20.5% (95% CI 18.7-22.3) infected by 2016 and 2017. Due to the limited temporal information provided by cases in this state, we assumed a constant FOI in Bahia between 2014-2017 to aid model convergence.

Maternally-derived antibodies can provide temporary protection in infants, but there are currently no studies quantifying the levels and duration of maternal antibodies against CHIKV. The seroprevalence and case data included in our analysis did not have enough granularity at an age level to robustly quantify the role of maternal antibodies in infections in infants. Consequently, we excluded all cases aged <1y from the fitting of our model. To nevertheless obtain estimates of the IFRs in individuals aged <1y, we computed the proportion of females between the ages of 16 and 45 that had been infected in each state and year, and assumed that an equivalent proportion of infants would be protected against CHIKV for six months in the following year. To also estimate the probability of detection in individuals aged <1y, we divided the number of observed cases in this age group by the number of infections inferred from <1-specific FOI.

### Sensitivity analyses

We investigated the probability that suspected cases are laboratory confirmed. We conducted a sensitivity analysis where, in addition to all previously mentioned data streams, we also incorporated annual aggregates of suspected chikungunya cases for each Brazilian state obtained from the Notifiable Diseases Information System (Sistema de Informação de Agravos de Notificação - SINAN). Importantly, this database and the GAL database, where the line list of laboratory confirmed cases was obtained from, are not cross-checked. This meant that for certain states, in certain years, there were more reported laboratory confirmed cases than suspected cases. Consequently, in this analysis the expected number of suspected diagnoses 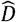 per cohort is determined by:

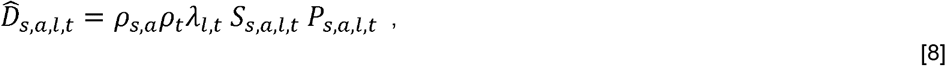

And the number of laboratory-confirmed cases is estimated using a state-specific probability of laboratory confirmation *Ψ*:

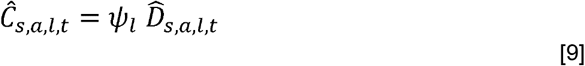

Finally, we fit the overall number of suspected diagnoses per state *D*_*l*_ assuming a Poisson distribution, in addition to the likelihoods specified in equations 7-9:

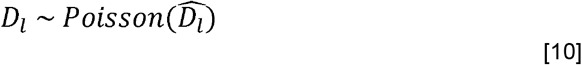

### Potential vaccine impact

We assumed that the vaccine can confer life-long protection against infection and against disease ^1^. Thus, susceptible vaccinated individuals have an annual probability of infection of (1 - vaccine efficacy against infection) * FOI, with an efficacy against infection of 70% in the base case scenario, consistent with previous modelling work ^1^. Unvaccinated infections have a probability of developing symptomatic disease of 50%, and the corresponding probability in vaccinated infections is (1 - vaccine efficacy against disease) / (1 - vaccine efficacy against infection) * 50%^25^. With a vaccine efficacy against disease assumed to be 95% in the base case scenario, vaccinated infections have a probability of 8.33% of developing symptomatic disease. We also measured the number of detected cases and deaths drawing detection probabilities and IFR samples from our model posterior. Additionally, we evaluated Disability-Adjusted Life Years (DALYs) using previously reported probabilities of occurrence, duration, and disability weights for acute, severe, and chronic disease following CHIKV infection.

We simulated CHIKV circulation over five years with and without vaccination, and measured potential vaccine impact in terms of infections, cases, detected cases, severe cases, deaths, and DALYs averted by the vaccine. For each metric, we reported the median estimated and 95% confidence intervals computed by running simulations over a bootstrap of 1,000 independent samples from our model posterior.

Additionally, we evaluated Disability-Adjusted Life Years (DALYs) averted from the vaccine, representing the number of healthy life years lost to disease or to premature death. DALYs are computed as the sum of Years Lived with a Disability (YLDs) and Years of Life Lost (YLLs). To calculate YLDs, we assumed that all cases have a probability of developing mild disease of 88% in the acute phase, with the remaining 12% having severe disease ^1^. Further, 40% of severe cases develop chronic arthralgia^25^. The acute phase of disease has a duration of seven days and that chronic phase of one year. The disability weights for each type of disease are 0.006 in mild acute cases, 0.133 in severe acute cases, and 0.233 in chronic cases ^51,52^. Separately, we compared national sex- and age-specific life expectancy estimates with the number of deaths in the corresponding group to compute the number of Years of Life Lost (YLLs).

